# Assessing the effectiveness of portable HEPA air cleaners for reducing particulate matter exposure in King County, Washington homeless shelters during the COVID-19 pandemic: implications for community congregate settings

**DOI:** 10.1101/2023.01.20.23284493

**Authors:** Ching-Hsuan Huang, Thu Bui, Daniel Hwang, Jeffry Shirai, Elena Austin, Martin Cohen, Timothy Gould, Timothy Larson, Igor Novosselov, Shirlee Tan, Edmund Seto

## Abstract

Over four thousand portable air cleaners (PACs) with high-efficiency particulate air (HEPA) filters were distributed by Public Health - Seattle & King County to homeless shelters during the COVID-19 pandemic. This study aimed to evaluate the real-world effectiveness of these HEPA PACs in reducing indoor particles and understand the factors that affect their use in homeless shelters. Four rooms across three homeless shelters with varying geographic locations and operating conditions were enrolled in this study. At each shelter, multiple PACs were deployed based on the room volume and PAC’s clean air delivery rate rating. The energy consumption of these PACs was measured using energy data loggers at 1-min intervals to allow tracking of their use and fan speed for three two-week sampling rounds, separated by single-week gaps, between February and April 2022. Total optical particle number concentration (OPNC) was measured at 2-min intervals at multiple indoor locations and an outdoor ambient location. The empirical indoor and outdoor total OPNC were compared for each site. Additionally, linear mixed-effects regression models (LMERs) were used to assess the relationship between PAC use time and indoor/outdoor total OPNC ratios (I/O_OPNC_). Based on the LMER models, one percent increase in the hourly, daily and total time PACs were used significantly reduced I/O_OPNC_ by 0.34 [95% CI: 0.28, 0.40], 0.51 [95% CI: 0.20, 0.78], 2.52 [95% CI: 1.50, 3.28], respectively, indicating that keeping PACs on resulted in significantly lower I/O_OPNC_ or relatively lower indoor total OPNC than outdoors. The survey suggested that keeping PACs on and running was the main challenge when operating them in shelters. These findings suggested that HEPA PACs were an effective short-term strategy to reduce indoor particle levels in community congregate living settings during non-wildfire seasons and the need for formulating practical guidance for using them in such an environment.

## 1. Introduction

Individuals experiencing homelessness account for a significant proportion of the US population, estimated at approximately 568,000 people each night in 2019, with the majority (63%) of homeless persons housed in shelters (Henry et al., 2020). Research has documented infectious disease outbreaks in homeless shelters, including airborne droplet transmission of *M. tuberculosis* in shelters operated in multiple U.S cities (Coffey et al., 2009; Martin et al., 2013, 2014). Findings from these previous studies included inspections of air handling and air flows and resulted in recommendations for the use of improved filtration, improved fresh air supplies, maintenance of existing ventilation units, and the need for written respiratory protection plans and separation of suspected infected individuals from the general population. With the COVID-19 pandemic, there has been renewed concern over the potential for airborne transmission of infectious droplets and particles in homeless shelters. Homeless people are more vulnerable to severe COVID-19 due to a higher burden of comorbidities, with estimates that they may be two to three times as likely to die of the disease than the general population (Culhane et al., 2020; Perri et al., 2020). These concerns have been partially supported by case reports of SARS-CoV-2 transmission in homeless shelters in different US metropolitan areas, including in King County, Washington (Baggett et al., 2020; Imbert et al., 2021; Mosites et al., 2020; Tobolowsky et al., 2020).

In King County, Washington, the single night count of individuals experiencing homelessness was estimated to be 13,368 in 2022, with 43% of the population sheltered (King County Regional Homelessness Authority, 2022). A case report from King County documented outbreaks in April 2020 at three homeless shelters, with 10.5% test positivity among the 181 residents and higher numbers of positives in the ensuing weeks afterward, including infections in both shelter occupants and staff members (Tobolowsky et al., 2020). In shelter settings, where masks and vaccinations are not consistently adopted, improving air quality may be one of the most effective interventions that can be deployed in congregate shelter settings to reduce SARS-CoV-2 transmission (Agarwal et al., 2021; Piscitelli et al., 2022).

Controlling infectious airborne droplets and particles in congregate living settings or homeless shelters is further complicated by the summer wildfire smoke season, which results in conflicting guidance on ventilation for indoor air. Generally, increasing ventilation and outdoor air exchange, and improving filtration may be considered for infection control. But, for managing wildfire smoke, it is recommended that outdoor air exchange be minimized to reduce the infiltration of outdoor smoke into the indoor environment. Managing the potential overlapping risks of SARS-CoV-2 transmission and wildfire smoke-related respiratory health effects may be especially challenging as there may be increased demand for and occupancy of homeless shelters (thus greater density of people) during wildfire smoke episodes (Seattle Human Services, 2021). Although generally less severe, a similar situation can occur in the winter during wood burning, which settles in the central low-lying areas of Seattle and King County, sometimes leading to poor air quality during winter inversion events. This could be a problem for congregate and emergency shelters that are set up during extreme weather events.

Portable air cleaners (PACs) equipped with a high-efficiency particulate air (HEPA) filter have been shown to be effective in reducing particle concentrations in several studies conducted in residential settings, and for wildfire smoke specifically. Multiple agencies, including the Centers for Disease Control (CDC) and the Environmental Protection Agency (EPA), have recommended using HEPA PACs to supplement HVAC systems to reduce indoor particle levels (Centers for Disease Control and Prevention, 2021). Barn et al. summarized some studies, many of which were based on randomized controlled study designs (Barn et al., 2016; U.S. Environmental Protection Agency, 2022), that support this recommendation. Henderson et al. documented up to 63-88% lowered PM_2.5_ concentrations with HEPA PACs (Henderson et al., 2005), while crossover studies by Barn et al. and Allen et al. found lower infiltration of smoke when PACs were used compared to when they were not (Allen et al., 2011; Barn et al., 2007). A recent study of HEPA PACs used during the September 2020 Washington State wildfire episode indicated PM2.5 reduction effectiveness ranged from 48-78% across seven homes (Xiang et al., 2021). Generally, these studies of PACs have been conducted in home settings rather than in community settings (e.g., clean air shelters in schools, libraries, or community centers) (Barn, 2014). Furthermore, despite the evidence supporting home HEPA PACs use for reducing particle exposure, there are challenges for PAC performance in multi-zone indoor environments. There remains considerable uncertainty in the performance of HEPA PACs in multi-zone congregate housing settings such as homeless shelters, where there may be competing decisions related to ventilation due to the need to manage both SARS-CoV-2 transmission and wildfire smoke. To date, there have been no studies presenting data on the real-world effectiveness of HEPA PACs for reducing particle exposures in larger multi-zone homeless shelters. Further, no empirical studies have quantified the usage of HEPA PACs in homeless shelters and attempted to correlate performance with site, building, or management decisions.

Since 2020, over 4,000 HEPA portable air cleaners were deployed at homeless shelters in King County, Washington, by Public Health – Seattle & King County (PHSKC) to help control the COVID-19 pandemic and protect the homeless population from acquiring infection. Considering the significant demand, shelters were prioritized for distribution using an equity tool that considered location, population served, and shelter resources. Multiple units were given to shelters for use in the common and sleeping areas. In this study, we aimed to evaluate the real-world effectiveness of these PACs in reducing indoor particles in these community congregate living settings. The objectives of this research were to understand the (1) usage pattern and (2) factors that affect the use of the HEPA PACs deployed at the shelters, and (3) the effectiveness of these PACs in reducing indoor particle levels, relative to the outdoor particle concentrations at each site.

## 2. Methods

### 2.1 Site selection and collection of site characteristics

Four rooms across three different homeless shelters (denoted as sites 1, 2a, 2b, and 3 hereafter) in King County, Washington with varying geographic locations and building/operating conditions were selected to participate in this study. These three sites were among the sites that were pre-selected by the county for HEPA PACs deployment. For each selected site, information on building openings (including doors and windows), operating schedules, HVAC system, floor plan, room size, and the primary indoor and outdoor particle sources were collected via field survey. Additional site characteristics, including the residential history of clients, were collected via a post-hoc survey. The survey was anonymous and administered to the site operators and clients aged 18 or older at the end of the study via email and paper. The survey also collected information about residents’ perceptions of air quality and pollution sources, and attitudes toward HEPA PACs. The study protocol and recruitment and consent procedures were approved by the University of Washington Human Subjects Division and the Washington State Institutional Review Board.

### 2.2 Deployment of HEPA PACs and usage monitoring

Multiple portable HEPA PACs (C535 3-stage True HEPA Air Purifier and XQ dual 4-stage True HEPA Air Purifier; Winix America) with brand new sets of filters were deployed in the sleeping dorm or main activity area of each shelter based on the room volume and the clean air delivery rate (CADR) rating of the PACs to achieve 5 air exchange rates per hour (Washington State Department of Health, 2022b). The locations of the PACs were recorded and tracked during the study. The C535 PACs contain three stages of filters, including a pre-filter, an activated carbon filter, and a HEPA filter. The XQ PACs contain two sets of 3-stage filters (a pre-filter, an activated carbon filter, and a HEPA filter) on the front and rear sides of the body. Both models of PAC contain a bipolar ionizer (which can be disabled) and provide five fan speed level settings, including sleep mode, fan speed 1 to 3, Turbo, and an “Auto-mode” feature (i.e., the fan speed level will be adjusted according to the feedback of the built-in air quality sensor). The detailed specifications of these two PACs and the measured energy consumption under different fan speed levels were summarized in Table A1. Before the second and third sampling round, the pre-filter of each PAC was vacuumed with a handheld vacuum cleaner to remove the dust built-up. The bipolar ionizer of each PAC was turned off before each sampling round.

The PACs were deployed and monitored for three two-week sampling rounds at each site, separated by single-week gaps, between February and April 2022. Each of the PACs deployed at each site was assigned a unique ID and plugged into a power data logger (HOBO® Plug Load Logger Model UX120-018; Onset Computer Corp.), which measured time-stamped energy usage at 1-minute intervals for the entire study period to allow tracking of their usage and fan speed. The logged data were downloaded by study staff for each data collection period. During the round 1 and round 2 deployments, the PACs were purposely set to operate on Auto-mode. During the round 3 deployment, the PACs were set to operate on fan speed level 3. However, the clients or shelter staff were allowed to change the fan speed setting however they wished during each deployment. At the beginning of each deployment, if a PAC was found unplugged or turned off, it was plugged back in and turned on according to the fan speeds noted above by the research staff.

### 2.3 Indoor and outdoor particle concentration monitoring

At each site, multiple indoor locations in the selected sleeping dorm or main activity area with PAC were monitored throughout the three sampling rounds using real-time air quality monitors (PurpleAir PA-II-SD; PurpleAir) placed and secured at the height of 1-2 m above the floor and at least 1 m away from any PAC or HVAC inlet/outlet. The PurpleAir PA-II-SD monitor contains two duplicate optical particle counters (OPC) (Plantower PMS 5003; Beijing Plantower Co. Ltd.), pressure, temperature, and humidity sensor (BME280; Bosch SensorTec). The OPC uses the laser scattering principle to measure the number of particles suspended in the air. The photodiode of the OPC is positioned perpendicular to the excitation beam and measures the ensemble scattering of particles in the optical volume. The measured scattering light intensity is converted to a voltage signal to estimate the number concentration of particles with an optical diameter ranging from 0.3 to 10 microns in six size bins (>0.3, > 0.5, >1.0, >2.5, >5.0, and >10.0 μm) and mass concentrations for PM_1_, PM_2.5,_ and PM_10_. In this study, the number concentration reported in size bin >0.3 μm was defined as the total optical particle concentration (OPNC). The data were timestamped and saved to the internal Secure Digital (SD) memory card at 2-minute intervals. Prior to their use in the study, these monitors were individually calibrated in a chamber experiment with woodsmoke particles against a real-time optical particle sizer (TSI optical particle sizer model 3330, TSI Inc.). The calibration shows R^2^ ranging from 0.97 to 0.99 for these monitors, and the root mean squared error (RMSE) of these calibrated monitors was less than 900 #/cm^3^ within the measurement range of 0 – 20000 #/cm^3^ (Table A2). The optical particle sizer was factory-calibrated prior to this study. In addition to the multiple indoor sampling locations at each site, a single PurpleAir PA-II-SD was placed outside at each shelter that monitored the outdoor ambient particle concentrations throughout the study periods. The outdoor locations were selected based on the representativeness of the general ambient air situation at each shelter, access to an electrical outlet, and were secured to minimize the potential for theft.

### 2.4 Statistical analysis

For the analysis, the 2-minute particle monitoring data were first aggregated hourly. The empirical indoor and outdoor total OPNC data were then compared within sites. Based on the Shapiro-Wilk tests, the indoor and outdoor total OPNC data were found not to be normally distributed. Thus, the Wilcoxon signed-rank tests (for paired comparison) were conducted to compare the indoor and outdoor total OPNC levels of each site.

Next, three PAC usage metrics were computed for each site: (1) the percent time the PACs were on; (2) the percent time the PACs were on different fan speed levels, including sleep mode, fan speed level 1 to level 3, and Turbo; and (3) the total power consumption of all PACs. Linear mixed effects regression (LMER), which incorporated random intercepts for sites to account for between-site correlations, as well as within-site correlations of repeated measurements, was used to examine the relationship between the indoor/outdoor total particle number concentration ratio (I/O_OPNC_) and different PAC usage metrics (Eq. (1) – (3)):

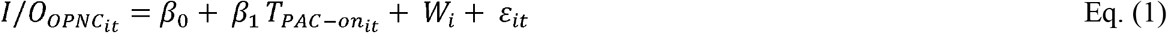

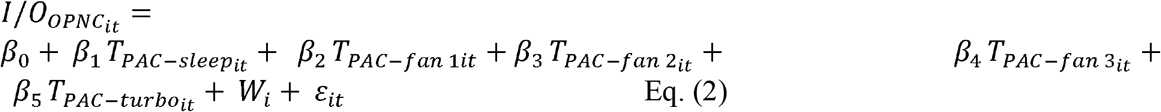

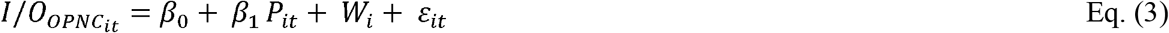

where 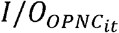 the indoor/outdoor total optical particle number concentration ratio of site i at time (hour) *t*; *β*_0_ – *β*_5_ are the coefficients of the LMER models; 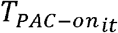 in Eq (1) is the percent time that the PACs were on of site i at time *t*, %; 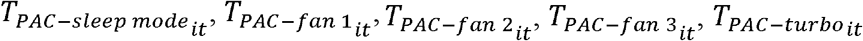 in Eq (2) are the percent time that the PACs were on sleep mode, fan speed 1, 2, 3, and Turbo of site i at time t, respectively, %; *P*_*it*_ in Eq (3) is the total power consumption of all PACs of site *i* at time *t*, Watts; *W*_*i*_ is the random effect factor, and *ε*_*it*_ is the residual. The LMER models were also assessed on daily and round levels. The outliers of 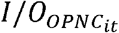 (i.e., measurements that were 1.5 interquartile range below the first quartile or above the third quartile) were removed for the modeling. For all statistical tests, p ≤ 0.05 indicated statistical significance in this study. All calculations and figures were made using “nlme”, “data.table”, and “ggplot2” packages in R Version 4.1.1 embedded in Rstudio Version 2021.09.0.

## 3. Results

### 3.1 Site characteristics

Table 1 summarizes the characteristics of the enrolled sites based on the field and post-hoc surveys. All three sites were located on the 1^st^ floor. Site 1 and site 3 were mechanically ventilated 24 hours per day with built-in HVAC systems, whereas site 2 (including two separate rooms 2a and 2b) was naturally ventilated without HVAC systems. Due to the study seasons (winter and spring), site 1 and site 3 used central heating systems to provide warmth to the rooms. Site 2 (including two separate rooms 2a and 2b) used wall radiators for heating. While windows were not available in the monitored area at site 1 and site 3, doors leading to the outdoor area were present and could have been opened during the study periods by shelter clients or staff. Site 1 is in the busy metro center and about 120 meters away from the major highway in the area. This site served approximately 20 clients from 9 am to 8 pm on weekdays, and 10 am to 2 pm on Saturdays. Site 2 (including two separate rooms 2a and 2b) is about 320 meters away from a major highway, whereas Site 3 is only 60 meters away from the closest highway. Site 2 (including two separate rooms 2a and 2b) and site 3 offered overnight services, were open 24 hours per day, seven days per week, and served approximately 50 and 100 clients per day, respectively. Onsite cooking took place only at sites 1 and 3, although already cooked meals were provided at sites 2a and 2b.

**Table 1.** General characteristics of the study sites.

| Site ID | Monitored area | Room size (m<sup>3</sup>) | Year first built | Ventilation type | Window opening | Cooking onsite |
| --- | --- | --- | --- | --- | --- | --- |
| Site 1 | Main activity area | 996 | 1922 | Mechanical | NA<sup>a</sup> | Y |
| Site 2a<sup>b</sup> | Sleeping dorm #1 | 921 | 1903 | Natural | Possible | N |
| Site 2b<sup>b</sup> | Sleeping dorm #2 | 1021 | 1903 | Natural | Possible | N |
| Site 3 | Sleeping dorm | 238 | 1975 | Mechanical | NA<sup>a</sup> | Y |

<sup>a</sup> No windows present in the monitored area.
<sup>b</sup> Site 2 had two areas monitored.

### 3.2 Measured PAC energy consumption and usage

Fig. 1 illustrates the measured % time of PACs operating under different fan speed levels at each site. The PACs deployed at site 1, site 2a, and site 2b were found operating under the Turbo fan speed during round 1 most of the time (∼45%). This could be because the shelter clients or staff adjusted the PAC fan speed settings manually to Turbo rather than keeping them running in Auto mode during the study. This assumption is supported by the time series of the individual PAC energy consumption shown in Appendix Fig. A.1-A.4. Fig. A.1 shows the time series of the 13 PACs deployed at site 1. During sampling round 1, the energy consumption of multiple PACs (e.g., PAC-003, PAC-004, and PAC-006) remained at ∼ 60 watts a majority of the time. This watt level corresponds to the energy consumption of the PAC running at Turbo fan speed (Table A.1). Even though the PACs were set to Auto-mode by the study staff at the beginning of sampling round 1, it is unlikely that the PACs remained at the Turbo fan speed for such extended periods of time due to elevated PM concentrations in the indoor environment. Similar PAC energy consumption can be observed for round 1 sampling at site 2a (Fig. A.2), site 2b (Fig. A.3), and site 3 (Fig. A.4). The total minutes of the PAC energy consumption monitored at each site are summarized in Table A.3. The incompleteness of site 1 and site 3 data was due to the power data loggers being unplugged from the PACs during the monitoring.

**Fig. 1.**
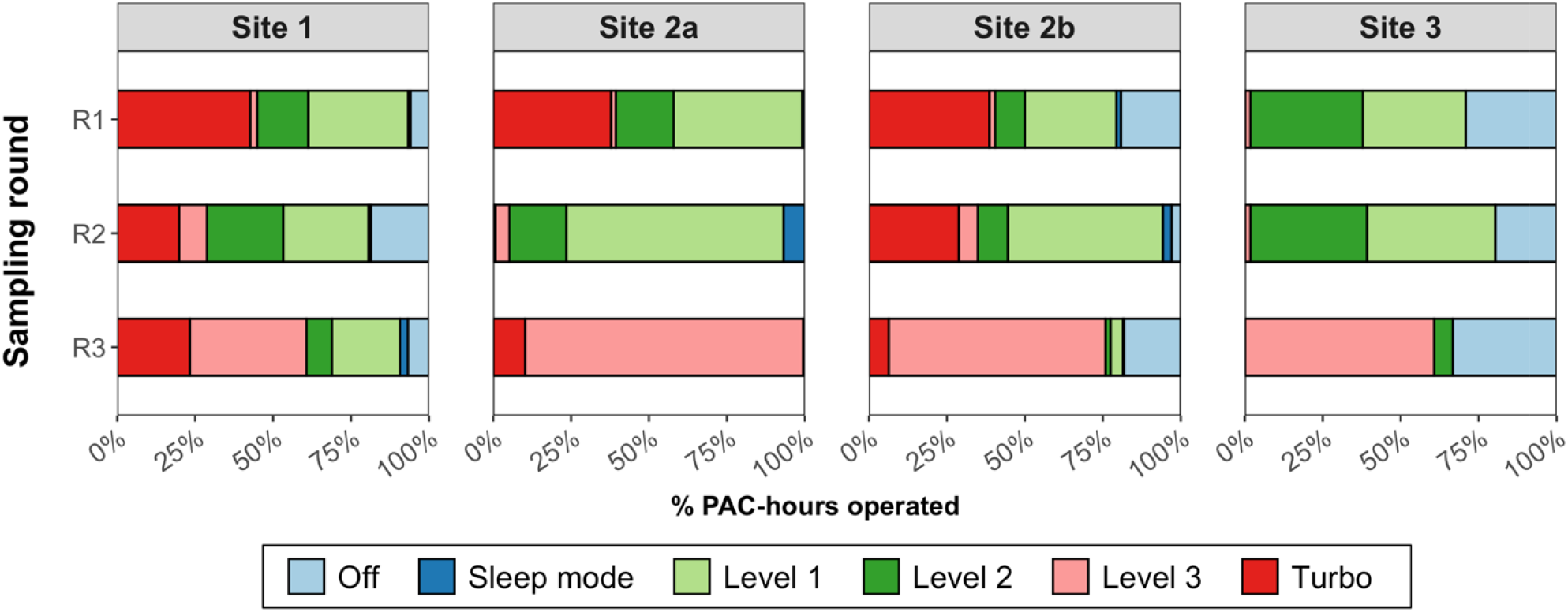
The measured % time of PACs operated under different fan speed levels. R1: round 1; R2: round 2; R3: round3. The PACs were initially set to operate on Auto-mode during round 1 and round 2, and fan speed level 3 during round 3.

### 3.3 Empirical particle concentrations

Table 2 shows the hourly mean indoor and outdoor total particle concentrations, and temperature and humidity at four sites. Overall, the mean indoor total OPNC concentrations at each site were relatively low during all sampling rounds (< 200 #/cm^3^). In contrast, the outdoor total OPNC concentrations were significantly higher than the indoor levels across all three sites and sampling rounds (p< 0.001). Pooling all sampling rounds together within each site, the mean (standard deviation, SD) reduction of hourly indoor total OPNC level was 47% (106%), 67% (73%), 32% (115%), and 66% (48%) compared to outdoor levels at site 1, site 2a, site 2b, and site 3, respectively. At site 1, multiple peaks were observed in the outdoor total OPNC levels, with a maximum concentration of 20298 #/cm^3^ during round 2 sampling (Table 2 & Fig. 2). Despite the significant difference between the mean indoor and outdoor total OPNC levels, the hourly indoor levels observed at each site sometimes were comparable to or higher than the outdoor levels (Fig. 2, site 2b). The heatmap in Fig. 3 shows the temporal variation in total indoor and outdoor OPNC levels. For site 1, over 90% of the indoor total OPNC observations were missing during round 1 sampling because the indoor monitors were unplugged by people at the site.

**Table 2.** Summary of the hourly averaged indoor and outdoor total particle concentration at three sites.

| Site | Round | Indoor total OPNC (#/cm<sup>3</sup>)<sup>a</sup> |  |  |  |  | Outdoor total OPNC (#/cm<sup>3</sup>) |  |  |  |  | p-value<sup>c</sup> |
| --- | --- | --- | --- | --- | --- | --- | --- | --- | --- | --- | --- | --- |
|  |  | Min | Median (IQR) | Mean (SD) | Max | N | Min | Median (IQR) | Mean (SD) | Max | N<sup>b</sup> |  |
| Site 1 | R1 | 0.50 | 94.43<br>(114.23) | 133.22<br>(124.36) | 750.89 | 659 | 91.26 | 609.39<br>(684.99) | 891.08<br>(1011.65) | 8882.23 | 327 | < 2.2e-16 |
|  | R2 | 6.63 | 101.28<br>(115.73) | 156.38<br>(169.90) | 1420.44 | 657 | 35.16 | 319.15<br>(344.75) | 780.26<br>(1982.68) | 20298.03 | 312 | < 2.2e-16 |
|  | R3 | 0.70 | 82.76<br>(95.01) | 89.85<br>(76.73) | 251.60 | 49 | 117.55 | 452.71<br>(321.34) | 639.17<br>(827.45) | 9850.99 | 276 | 1.82e-12 |
| Site 2a | R1 | 0.51 | 53.22<br>(64.10) | 64.51<br>(52.35) | 346.7 | 610 | 31.61 | 286.26<br>(274.20) | 346.28<br>(237.02) | 1537.67 | 329 | < 2.2e-16 |
|  | R2 | 0.11 | 61.02<br>(58.78) | 78.18<br>(80.97) | 639.93 | 652 | 13.19 | 427.94<br>(349.83) | 452.46<br>(270.22) | 1983.50 | 330 | < 2.2e-16 |
|  | R3 | 0.27 | 53.67<br>(57.67) | 70.41<br>(82.45) | 636.80 | 619 | 8.05 | 289.38<br>(339.51) | 357.79<br>(243.59) | 1259.93 | 330 | < 2.2e-16 |
| Site 2b | R1 | 3.60 | 101.57<br>(99.47) | 163.91<br>(264.25) | 2768.55 | 637 | 31.61 | 286.26<br>(274.20) | 346.28<br>(237.02) | 1537.67 | 329 | < 2.2e-16 |
|  | R2 | 4.82 | 102.39<br>(92.41) | 131.16<br>(109.34) | 999.80 | 659 | 13.19 | 427.94<br>(349.83) | 452.46<br>(270.22) | 1983.50 | 330 | < 2.2e-16 |
|  | R3 | 0.42 | 122.88<br>(143.37) | 161.51<br>(134.86) | 980.24 | 660 | 8.05 | 289.38<br>(339.51) | 357.79<br>(243.59) | 1259.93 | 330 | < 2.2e-16 |
| Site 3 | R1 | 14.40 | 77.15<br>(71.87) | 99.82<br>(72.38) | 431.55 | 330 | 21.40 | 376.63<br>(291.69) | 433.78<br>(275.73) | 1574.26 | 329 | < 2.2e-16 |
|  | R2 | 6.60 | 44.89<br>(31.92) | 49.44<br>(33.16) | 337.44 | 330 | 44.11 | 286.86<br>(278.43) | 318.00<br>(194.83) | 1165.74 | 330 | < 2.2e-16 |
|  | R3 | 8.87 | 64.19<br>(54.19) | 85.12<br>(86.86) | 1098.08 | 330 | 43.49 | 275.72<br>(208.92) | 283.39<br>(159.06) | 1148.29 | 330 | < 2.2e-16 |

<sup>a</sup> For sites with multiple indoor monitors (site 1, site 2a, and site 2b), the average concentrations across all indoor monitors were presented.
<sup>b</sup> Number of data points.
<sup>c</sup> Comparison between the indoor and outdoor total OPNC based on the Wilcoxon signed-rank tests (for paired comparison).

**Fig. 2.**
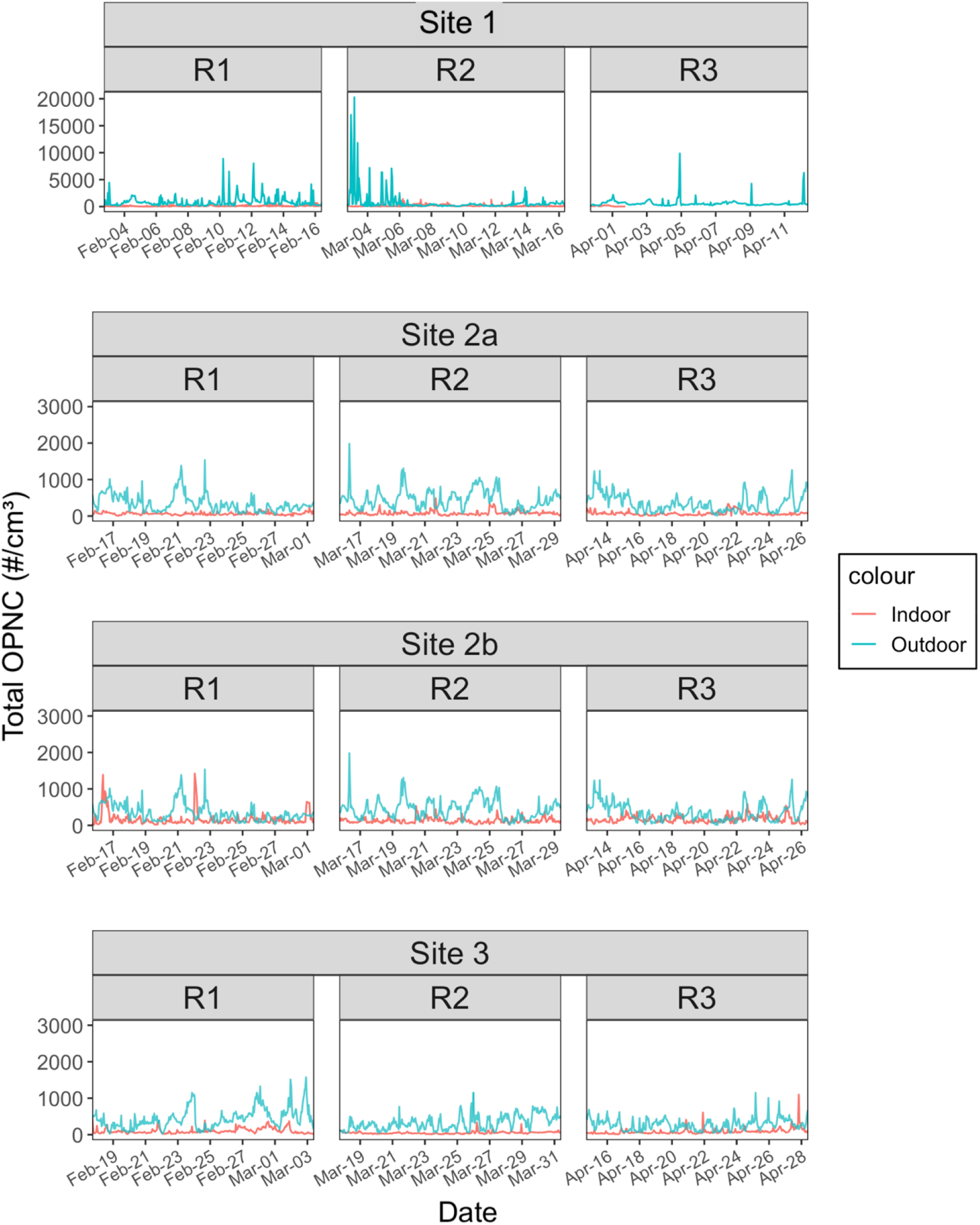
Time series of the indoor and outdoor total OPNC at three sites. For sites with multiple indoor monitors (site 1, site 2a, and site 2b), the average concentrations were plotted. The y-axis of site 1 was plotted on a different scale.

**Fig. 3.**
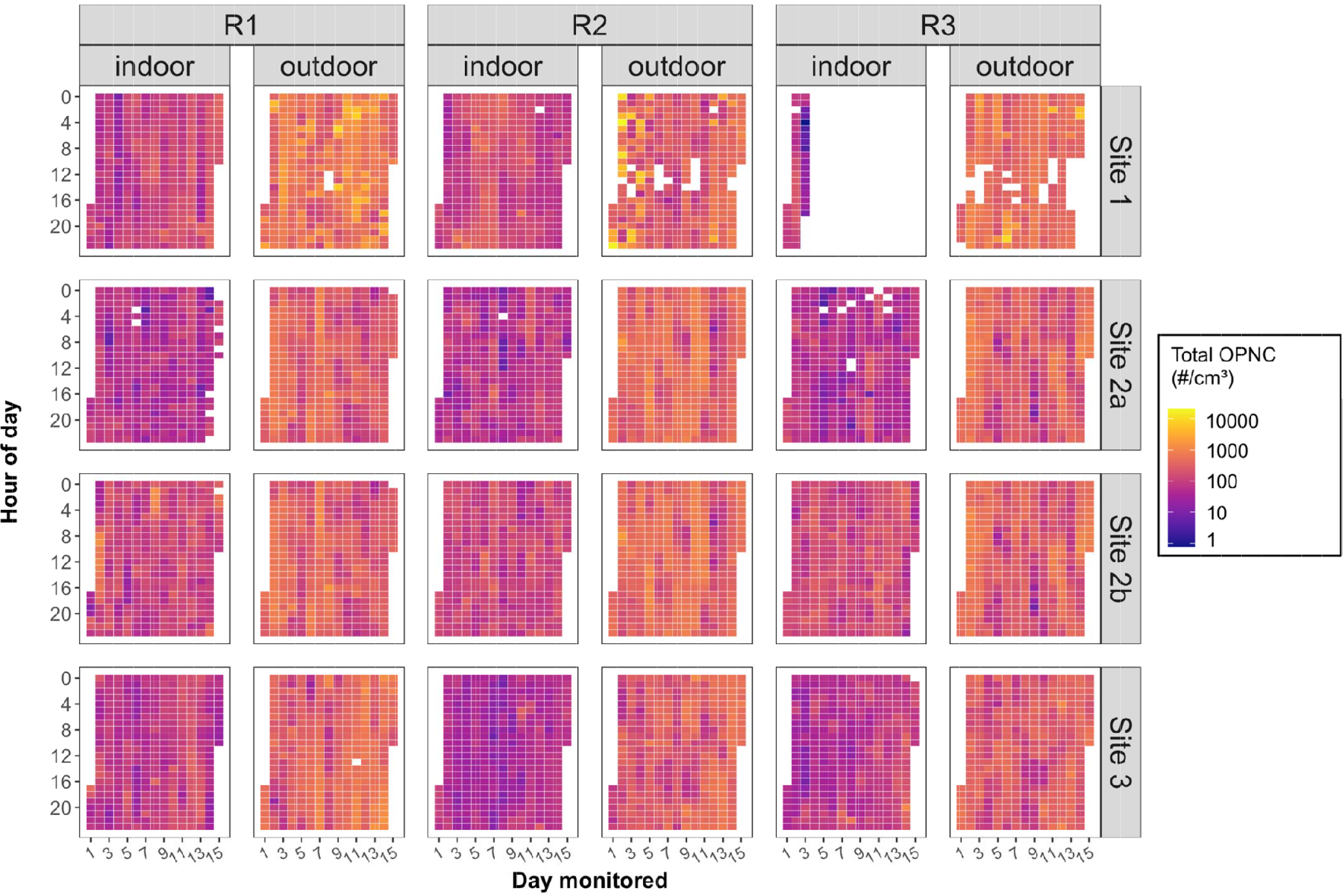
Heatmap of the indoor and outdoor total OPNC at each site. For sites with multiple indoor monitors (site 1, site 2a, and site 2b), the average concentrations were plotted.

Fig. 4 and Table 3 show the hourly indoor/outdoor total OPNC ratios (I/O_OPNC_) under different sampling rounds at each site. For all sites, the mean hourly I/O_OPNC_ during all sampling rounds was lower than 1, indicating that the indoor total OPNC levels were lower than the outdoor ones. However, the maximum I/O ratio at each site was larger than 1 (the data for round 3 of site 1 was excluded from the discussion due to incompleteness), suggesting the presence of indoor particle sources at each site.

**Fig. 4.**
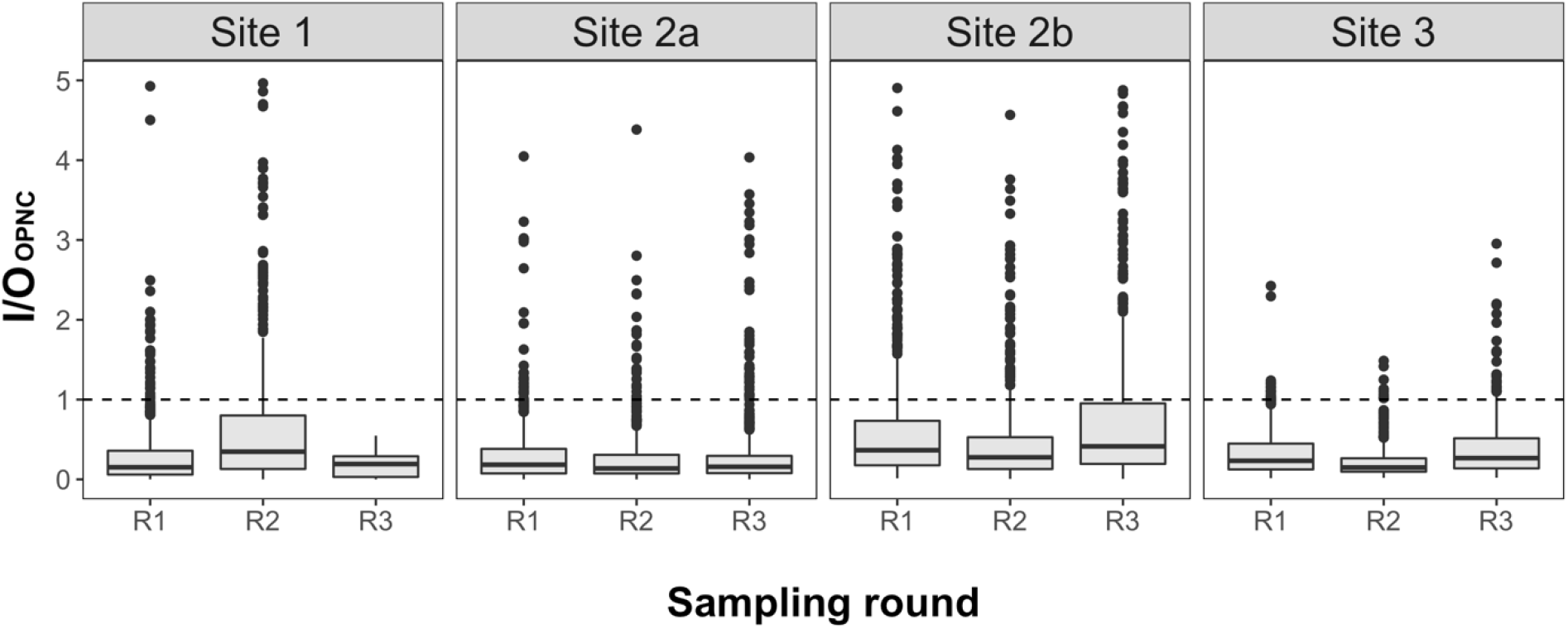
Boxplot of the hourly averaged indoor/outdoor particle number concentration ratio (I/O_OPNC_). Data points larger than 5 were excluded from plotting (max values are provided in Table 3).

**Table 3.** Summary of the hourly averaged indoor/outdoor total particle concentration ratio (I/O_OPNC_) at three sites.

| Site | Round | $I/O_{OPNC}$ | | | | $N^a$ |
| --- | --- | --- | --- | --- | --- | --- |
|  |  | Min | Median (IQR) | Mean (SD) | Max |  |
| Site 1 | R1 | <0.01 | 0.15<br>(0.30) | 0.29<br>(0.43) | 4.93 | 653 |
|  | R2 | <0.01 | 0.37<br>(0.72) | 0.79<br>(1.43) | 19.45 | 623 |
|  | R3 | <0.01 | 0.19<br>(0.26) | 0.18<br>(0.15) | 0.55 | 40 |
| Site 2a | R1 | <0.01 | 0.18<br>(0.31) | 0.31<br>(0.42) | 4.05 | 610 |
|  | R2 | <0.01 | 0.14<br>(0.24) | 0.28<br>(0.43) | 8.10 | 652 |
|  | R3 | <0.01 | 0.16<br>(0.23) | 0.39<br>(1.11) | 18.82 | 619 |
| Site 2b | R1 | 0.01 | 0.37<br>(0.58) | 0.71<br>(1.27) | 16.50 | 635 |
|  | R2 | 0.01 | 0.28<br>(0.41) | 0.49<br>(0.73) | 8.10 | 659 |
|  | R3 | 0.01 | 0.42<br>(0.79) | 0.84<br>(1.34) | 18.98 | 660 |
| Site 3 | R1 | 0.01 | 0.23<br>(0.32) | 0.33<br>(0.31) | 2.42 | 329 |
|  | R2 | 0.02 | 0.15<br>(0.17) | 0.23<br>(0.23) | 1.49 | 330 |
|  | R3 | 0.02 | 0.27<br>(0.38) | 0.46<br>(0.73) | 8.52 | 330 |

<sup>a</sup> The number of complete hours with both indoor and outdoor measurements.

### 3.4 Relationship between PAC usage metrics and indoor/outdoor total OPNC ratio (I/O_OPNC_)

Table 4 presents the results of the LMER models. Models 1-3 assessed the relationship between the I/O_OPNC_ and the “percent time the PACs were on,” “percent time the PACs were on different fan speed levels,” and “hourly total power consumption of all PACs” metrics, respectively, on different time averaging scales. Regardless of the time averaging scale, the regression coefficients were negative for the percent time PACs were on (β_1_) in model 1, indicating that keeping PACs on resulted in significantly lower I/O_OPNC_ or relatively lower indoor total OPNC than outdoors. One percent increase in the hourly, daily and total time PACs were used significantly reduced I/O_OPNC_ by 0.34 [95% CI: 0.28, 0.40], 0.51 [95% CI: 0.20, 0.78], 2.52 [95% CI: 1.50, 3.28], respectively. Fig. 5 shows the predicted hourly I/O_OPNC_ under 50% to 100% of PACs operating time, based on model 1 with an hourly averaging scale. Overall, hourly PAC operating time ranging from 50% to 100% results in I/O_OPNC_ smaller than 1, and with the increasing amount of hourly PAC operating time, the I/O_OPNC_ becomes lower.

**Table 4.** Summary of the results for the linear mixed-effects regression (LMER) models.

| Model | Averaging Scale | Coefficient <sup>a</sup> | Estimate | Standard error | 95% CI | R<sup>2</sup> |
| --- | --- | --- | --- | --- | --- | --- |
| Model 1 | Hourly | $\beta_0$ | 0.59 | 0.04 | 0.53 to 0.67 | 0.07 |
| | | $\beta_1$ | -0.34 | 0.03 | -0.40 to -0.28 | |
| | Daily | $\beta_0$ | 0.79 | 0.13 | 0.51 to 1.04 | 0.18 |
| | | $\beta_1$ | -0.51 | 0.14 | -0.78 to -0.20 | |
| | Round | $\beta_0$ | 2.58 | 0.40 | 1.69 to 3.34 | 0.96 |
| | | $\beta_1$ | -2.52 | 0.39 | -3.28 to -1.50 | |
| Model 2 | Hourly | $\beta_0$ | 0.60 | 0.04 | 0.53 to 0.67 | 0.07 |
| | | $\beta_1$ | -0.15 | 0.10 | -0.34 to 0.05 | |
| | | $\beta_2$ | -0.35 | 0.03 | -0.41 to -0.28 | |
| | | $\beta_3$ | -0.36 | 0.04 | -0.43 to -0.29 | |
| | | $\beta_4$ | -0.36 | 0.04 | -0.43 to -0.28 | |
| | | $\beta_5$ | -0.32 | 0.04 | -0.40 to -0.23 | |
| | Daily | $\beta_0$ | 0.70 | 0.14 | 0.44 to 0.98 | 0.26 |
| | | $\beta_1$ | -1.58 | 0.99 | -3.45 to 0.36 | |
| | | $\beta_2$ | -0.60 | 0.16 | -0.91 to -0.29 | |
| | | $\beta_3$ | -0.52 | 0.25 | -1.03 to -0.04 | |
| | | $\beta_4$ | -0.46 | 0.17 | -0.79 to -0.15 | |
| | | $\beta_5$ | -0.05 | 0.19 | -0.45 to 0.30 | |
| | Round | $\beta_0$ | 3.12 | 0.41 | 2.39 to 3.90 | 0.99 |
| | | $\beta_1$ | -18.79 | 4.60 | -25.56 to -11.97 | |
| | | $\beta_2$ | -1.25 | 0.50 | -1.98 to -0.52 | |
| | | $\beta_3$ | -4.62 | 0.87 | -5.90 to -3.33 | |
| | | $\beta_4$ | -2.97 | 0.30 | -3.42 to -2.53 | |
| | | $\beta_5$ | -4.05 | 0.50 | -4.79 to -3.29 | |
| Model 3 | Hourly | $\beta_0$ | 0.30 | 0.01 | 0.28 to 0.32 | 0.01 |
| | | $\beta_1$ | 1.70e-04 | 1.17e-04 | -5.29e-05 to 4.10e-04 | |
| | Daily | $\beta_0$ | 0.34 | 0.03 | 0.30 to 0.38 | 0.02 |
| | | $\beta_1$ | 4.41e-04 | 9.87e-04 | -1.14e-03 to 2.57e-03 | |
| | Round | $\beta_0$ | 0.48 | 0.09 | 0.32 to 0.65 | 0.12 |
| | | $\beta_1$ | -3.95e-03 | 0.01 | -0.01 to 0.01 | |

**Fig. 5.**
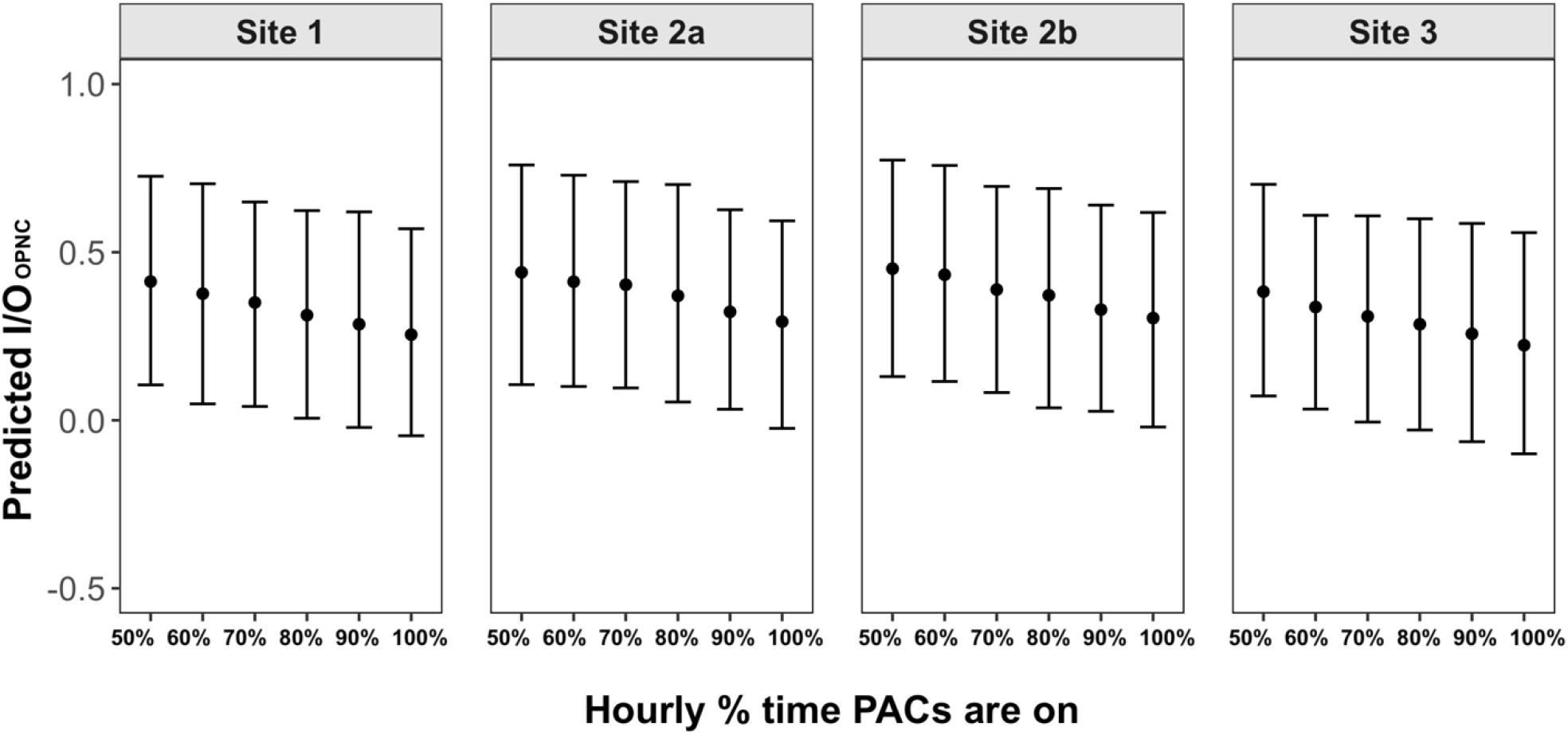
Prediction of hourly I/O_OPNC_ under 50% to 100% of hourly PACs using time at each site. The middle point represents the mean, and the top and bottom bars represent the upper and lower 95% confidence interval, respectively.

Similarly, in model 2 the correlation coefficients for the percent time PACs were on sleep mode (β_1_), fan speed level 1 (β_2_), level 2 (β_3_), level 3 (β_4_), and Turbo (β_5_) were negative. However, the reductions in these regression coefficients were non-linear, indicating that running the PACs at higher fan speed did not result in lower I/O_OPNC_ than at lower fan speed. One percent increase in the total time PACs running at sleep mode, fan speed level 1, level 2, level 3, and Turbo significantly reduced I/O_OPNC_ by 18.79 [95% CI: 11.97, 25.56], 1.25 [95% CI: 0.52, 1.98], 4.62 [95% CI: 3.33, 5.90], 2.97 [95% CI: 2.53, 3.42], and 4.05 [95% CI: 3.29, 4.79], respectively. The coefficient of determination (R^2^) of model 1 and model 2 with different time averaging scales ranging from 0.07 to 0.99, meaning that these LMER models can explain 7% - 99% of the data variance, with the 2-week models having the highest R^2^.

In model 3 with different time averaging scales, the regression coefficients of the total power consumption of all PACs (β_1_) were negative but not significant. One wattage increase in total PACs energy consumption results in an insignificant reduction of 0.004 [95% CI: -0.01, 0.01] of I/O_OPNC_, meaning that higher total energy consumption of PACs did not significantly lower I/O_OPNC_. The R^2^ values of model 3 were relatively low compared to model 1 and model 2. The highest R^2^ was the model with a 2-week time averaging scale (R^2^ = 0.12), indicating this model can only explain 12% of data variance.

### 3.5 Survey results

A total of 10 clients and 12 staff from three sites participated in the post-hoc survey. Most respondents reported that cooking fumes (17, 77.3%) and cigarette smoke outdoors (17, 77.3%) were the main indoor sources of air pollution. Body and bathroom odors (2, 9.1%), vehicle exhaust (2, 9.1%), and indoor vaping (1, 4.5%) were also reported as indoor sources of air pollution. Of 22 respondents, 16 (72.7%) felt air quality was better with PACs. Among the respondents who did not feel air quality was better with PACs (4, 18.2%, 2 missing), three respondents were clients. These clients reported their ability to smell air fresheners, cigarette smoke, cooking fumes, and vehicle exhaust as the reasons they didn’t feel the air quality was better. The only staff reported that air quality was not better with the PACs indicated it was due to continued smells of cooking fumes.

Regarding the maintenance and operations of PACs, half (11, 50%) of the respondents reported they hardly noticed staff cleaning PACs. Nearly 42% (5) of the staff responded that keeping the PACs running and the noise were the two primary concerns of operating the PACs. Among nine clients who responded to this question, over half (6, 66.7%) responded that they slept better with the PACs on and when the air quality was better.

## 4. Discussion

To our knowledge, this study is the first to examine the use of portable HEPA air cleaners and their impacts on indoor total particle concentration in homeless shelters. The results of this study suggest that using a sufficient number (estimated to achieve 5 air changes per hour based on the room size) of HEPA PACs and increasing the amount of time they are turned on, can significantly reduce indoor total OPNC compared to the outdoors, in real-world operating fan speeds of the air cleaners at homeless shelters.

A congregate living setting, by definition, is a facility or housing where people reside and share at least one common room, such as a sleeping room, bathroom, or kitchen. In homeless shelters where supportive services such as meals and housekeeping are provided, various sources of particles could have existed, such as cooking fumes, the use of vacuums, and air freshener. In this study, though PACs were found to reduce indoor total OPNC, elevated peaks (> 250 #/cm^3^) were still observed at each site. This highlighted the importance of source control. For example, staff and clients from site 1 and site 3 reported cooking fumes as the major indoor air pollution source, and clients from site 3 specifically mentioned that cigarette smoke from outdoors could be smelled in the sleeping dorm. This could explain the indoor total OPNC peaks observed at these two sites (Table 2). At site 1 where the highest outdoor total OPNC was observed (20298 #/cm^3^), the staff reported smoking activities happened right next to the location of outdoor particle sensors during round 2 sampling (Table 2). In addition to these common indoor particle sources, wildfire smoke is a concern generally for our region, and outdoor regional particle concentrations can reach high concentrations in the late summer-fall season. Wildfire smoke may not have been reported because of the timing of our survey which was conducted in the spring season.

Our study results show that with the use of PACs, the empirical indoor total OPNC level was reduced by up to 67% compared to the outdoor levels. The LMER results of models 1 and 2 also supported that using PACs would result in lower indoor/outdoor total OPNC ratios. In model 3, the trend of regression coefficients (β_1_) of model 3 was the same (i.e., negative) as those of model 1 and model 2 (i.e., higher PACs energy consumption results in lower indoor/outdoor total OPNC ratios). However, these coefficients were not statistically significant. This might be due to noise in the data, or the variations in the energy consumption of the PACs operating at different fan speed levels were too small for the regression model to pin down the relationship between PAC energy consumption and indoor/outdoor total OPNC ratio. The energy consumption of the PACs operating at lower fan speed levels, including sleep mode, fan speed levels 1, 2, and 3 ranges from 2.8 – 10.4 and 8.7 – 21.9 wattages for the two models used in this study (Table A1). Nevertheless, the results of model 1 and model 2 concluded that the amount of time using the PACs was associated with lower indoor/outdoor total OPNC ratios.

Care should be taken when choosing which PACs for use in homeless shelters. Consistent with the existing guidance (Centers for Disease Control and Prevention, 2021; Public Health - Seattle & King County, 2021; The American Society of Heating, 2021; U.S. Environmental Protection Agency, 2022; Washington State Department of Health, 2022a, 2022b), this study suggested selecting HEPA PACs based on the CADR ratings and recommended working room size. While the existing guidance is formulated based on the CADR and room size at the highest fan speed settings (provided by manufacturers), the results of this study suggested that keeping HEPA PACs on all the time could significantly reduce indoor particle levels, regardless of what fan speed they are on. According to the survey results, the main challenge of operating the PACs on site reported by the shelter staff was to keep them on and running. For example, staff from the participating sites reported that PACs were unplugged from the electrical outlets to plug in other or personal electronic devices. In light of this, electrical outlets should be secured to prevent PACs from being unplugged. The use of labels or signage to explain the purpose of the PACs and communicating with staff and clients to keep them plugged in or turned on would also help address this challenge. Other recommendations in the existing guidance include selecting HEPA PACs with third-party verification (e.g., California Air Resources Board (CARB) and Association of Home Appliance Manufacturers (AHAM) certification) to avoid devices that could emit harmful gas (e.g., ozone) emissions. In the current study, noise was another concern that staff voiced regarding PAC use at shelter sites. There is growing evidence suggesting that noise is a major factor that affects behavior or attitude toward using PACs (Brugge et al., 2013; Huang et al., 2021). Therefore, noise level should also be considered when selecting PACs to use. Using lower settings can reduce the noise levels, so additional PACs can be helpful to provide more air changes per hour if used in a setting where the highest fan speed setting is not practical.

The existing guidance rarely discussed the costs of running and maintaining PACs. Based on the energy consumption data this study collected, the average monthly energy consumption per PAC ranges from ∼3.4 kWh to ∼17.4 kWh depending on the PAC model used and the percent time PAC operating under different fan speed levels. Other considerations include the cost and frequency of filter replacement, the need to designate staff that can clean and maintain PACs, and the design and ease of using the PACs.

This study has several limitations. First, the present study was not conducted during the wildfire season when the indoor particle concentrations would likely be higher. There may have been seasonal effects depending on the air pollution experienced at different times of the year.

Therefore, the effectiveness of PACs in lowering indoor particle concentration in wildfire seasons might be different. Second, detailed time-activity information was not collected in this study, which limits our ability to characterize the impacts of sources (e.g., cooking, cleaning, window opening, etc.) on indoor particle levels. Third, compared to previous studies that relied on cross-over design, this study was observational because the air cleaners were deployed not for the study but for COVID-19 control; it would have been unethical to randomize air cleaner use or sham filtration in this situation. Fourth, the applicability of the study results might be limited. Homeless shelters are not the sole type of congregate living setting. Some other common congregate living settings include nursing homes and correctional facilities. These settings could have different characteristics such as building conditions, the density of persons, the amount of time that people share a common space, and the sources of air pollution exposures. For facilities that open overnight, the concern about using PACs might be different (e.g., noise from PACs could be an issue in a sleeping dorm). Lastly, this study did not measure air exchange rates directly, which could impact the infiltration of particles of outdoor origins (Xiang et al., 2021; Zauli-Sajani et al., 2018). This may lead to a biased comparison between the indoor and outdoor particle concentrations and the calculation of indoor/outdoor particle concentration ratios. As previously mentioned, air filtration with PACs is recommended as a supplement to ventilation by various agencies, and there has been a growing interest in comparing the combined effectiveness of various ventilation and air filtration strategies. The objective of this study was not to answer such questions. Instead, our goal was to investigate the real-world effectiveness of HEPA PACs in reducing indoor particle concentration and provide qualitative insights on what factors impact the use of HEPA PACs in congregate living settings. Future studies building upon the current research and examining the effectiveness of HEPA PACs during different exposure scenarios (e.g., wildfire season) are warranted to formulate more comprehensive guidance for reducing indoor exposures. However, as discussed, our study findings suggest that short-term use of HEPA PACs is effective in reducing indoor particle levels in community congregate settings.

## 5. Conclusions

This study shows that portable HEPA air cleaners are an effective short-term strategy to reduce indoor particle levels in community congregate settings during non-wildfire seasons, though the overall effectiveness depended on the length of time that the portable HEPA air cleaners were used. Keeping portable HEPA air cleaners on and running was the main challenge when operating them in shelters. These findings suggested the need for formulating practical guidance for using portable HEPA air cleaners in community congregate settings.

## Data Availability

Data not available due to participant consent.

## Acknowledgment

The authors wish to express special thanks to King County Health Engagement Action Resource Team (HEART Team) for mobilizing rapid HEPA air cleaner deployment and facilitating connection with study sites, and PHSKC Environmental Health Services COVID Recovery Program on Indoor Air for assisting with logistics and resources.

