## Supplementary material for "Assessing the effectiveness of portable HEPA air cleaners for reducing particulate matter exposure in King County, Washington homeless shelters during the COVID-19 pandemic: implications for community congregate settings"

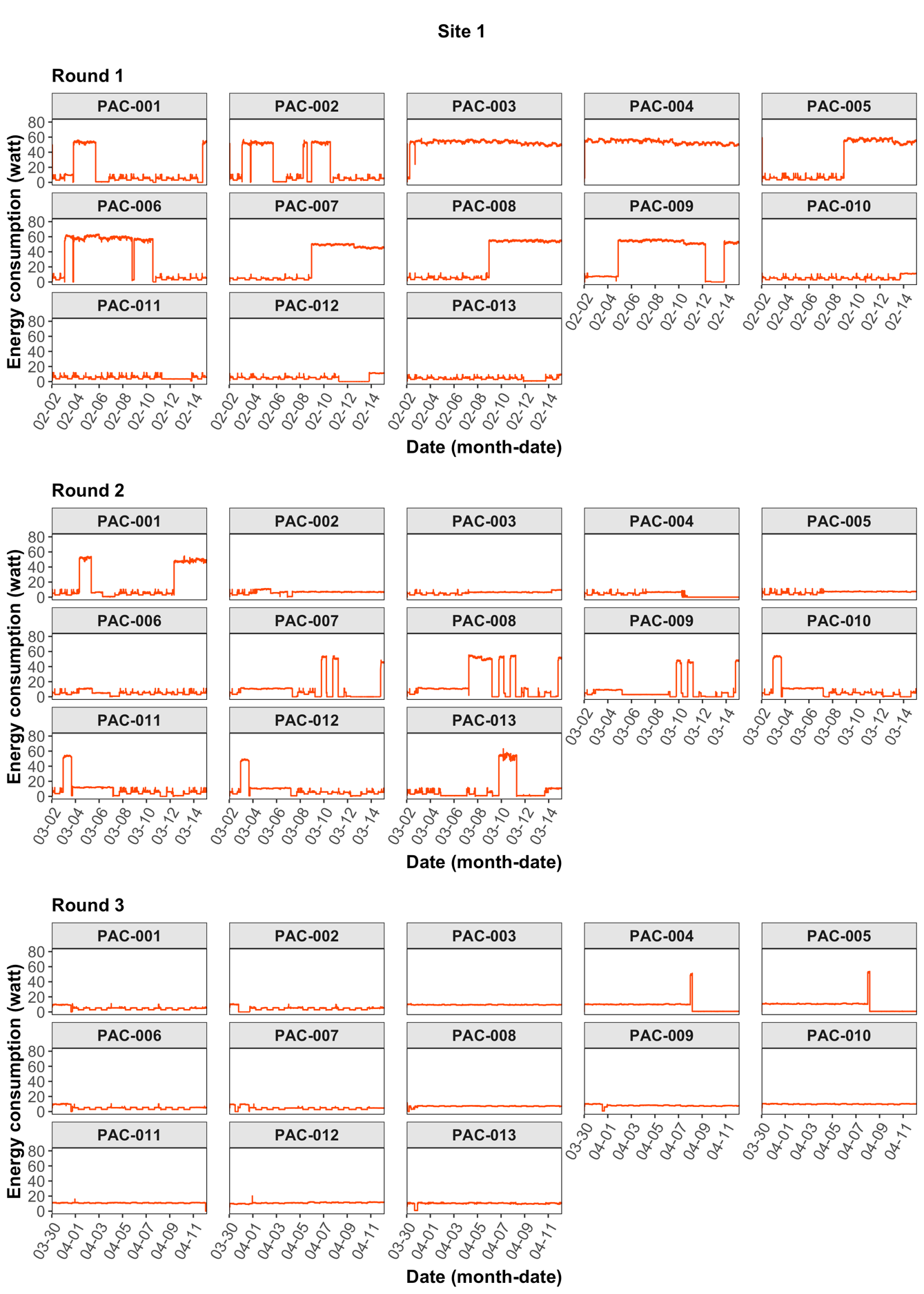

**Figure A1. Time series of the energy consumption of the PACs deployed at site 1 during three sampling rounds.**

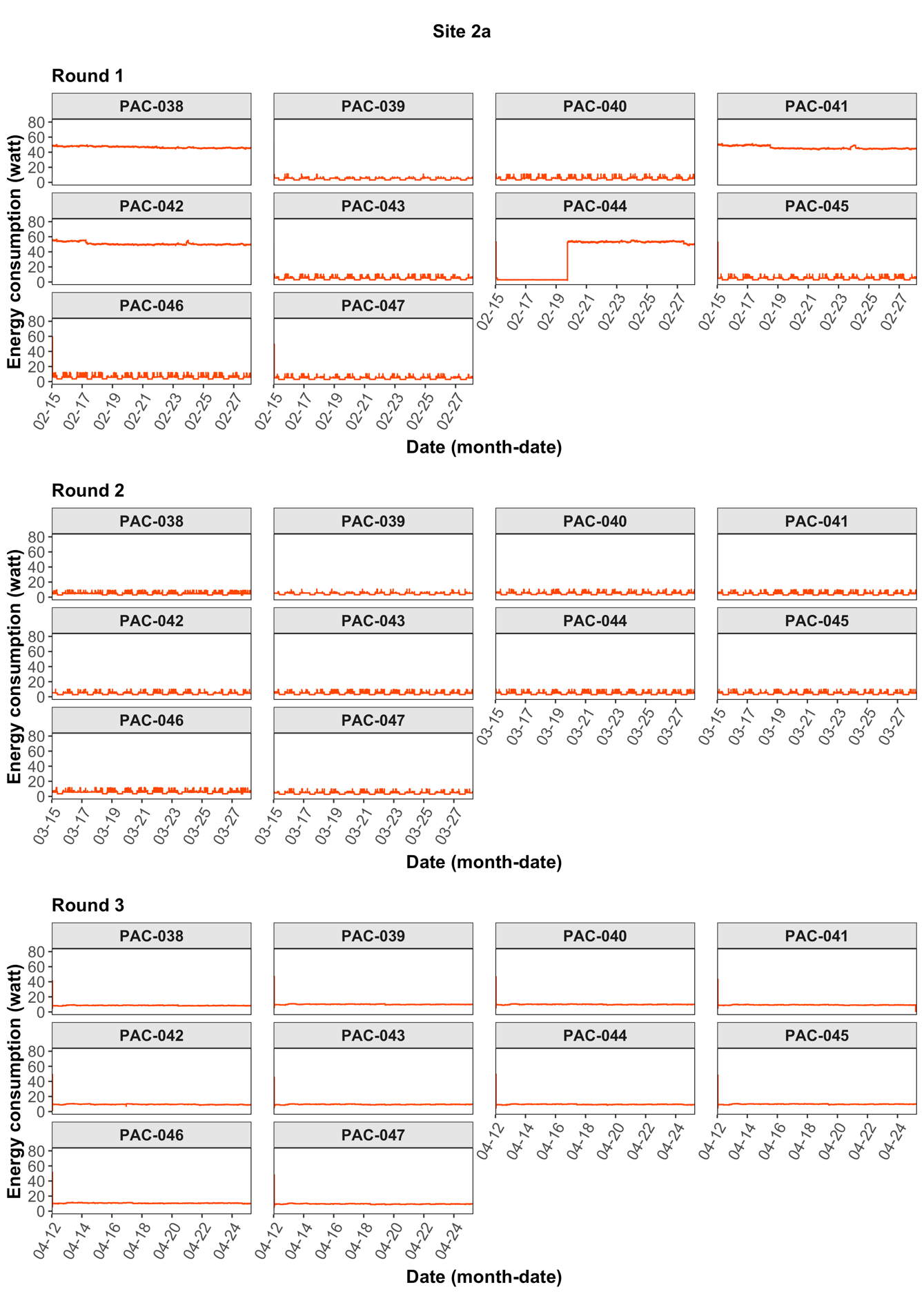

**Figure A2. Time series of the energy consumption of the PACs deployed at site 2a during three sampling rounds.**

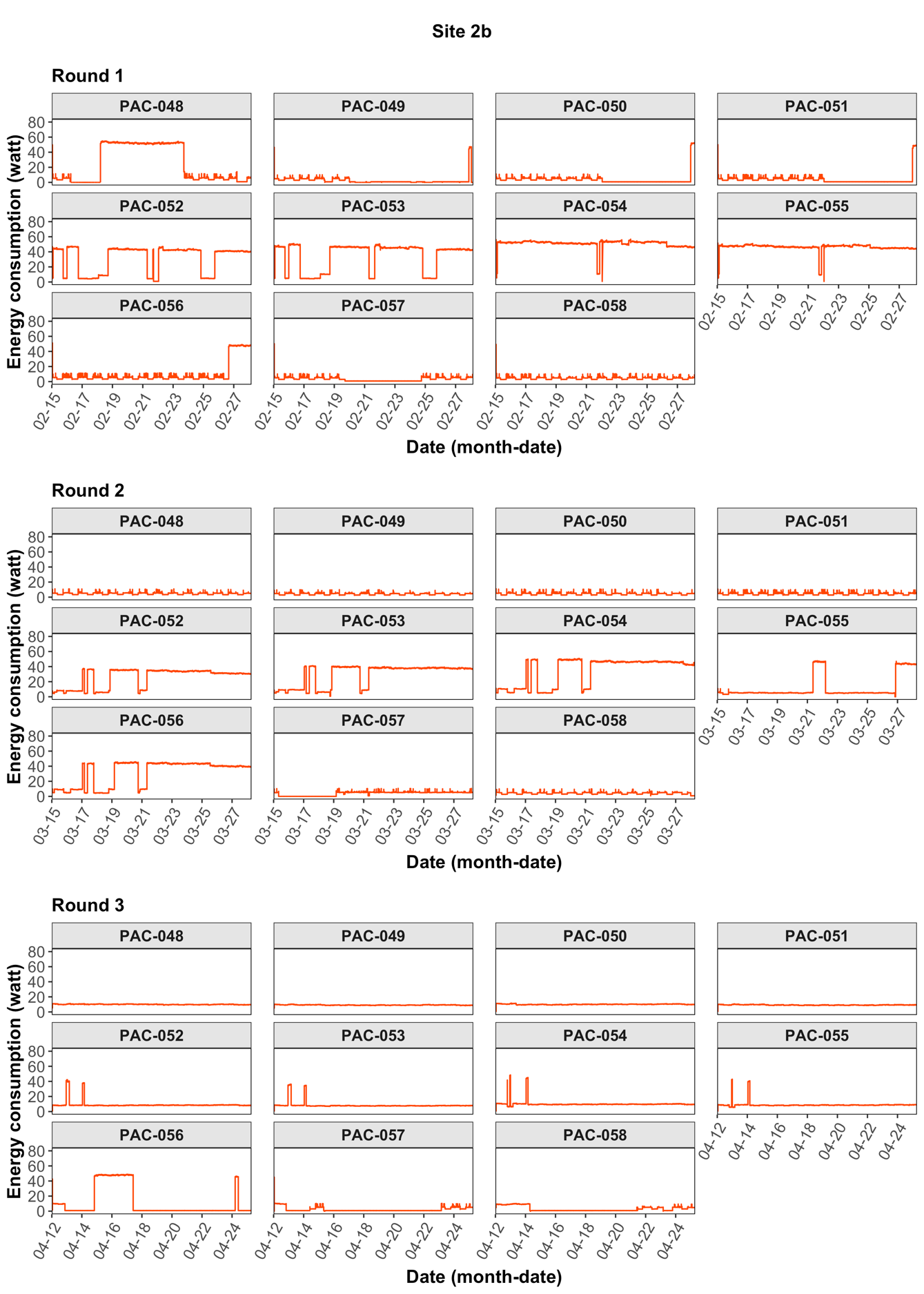

**Figure A3. Time series of the energy consumption of the PACs deployed at site 2b during three sampling rounds.**

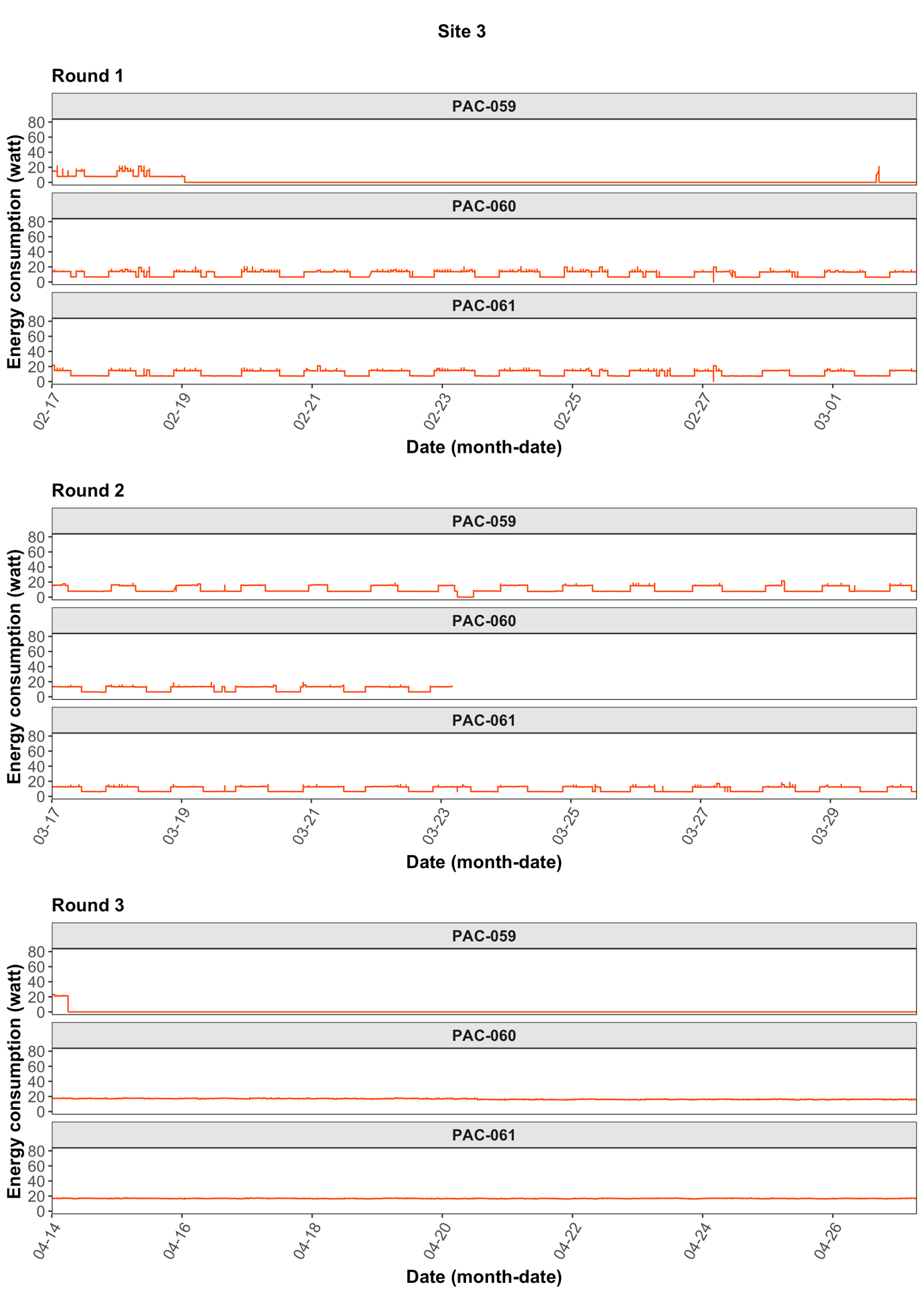

**Figure A4. Time series of the energy consumption of the PACs deployed at site 3 during three sampling rounds.**

**Table A1. Specifications of the HEPA PACs used in this study.**

| **Model** | **Winix C535** | **Winix Tower XQ** |
| --- | --- | --- |
| CADR (dust/pollen/smoke) ^a^ | 243/236/242 | 360 / 405 / 419 |
| Energy consumption (watt) ^b^ |  | |
| Speed sleep mode | 2.8 | 8.7 |
| Speed level 1 | 5.4 | 15.6 |
| Speed level 2 | 7.3 | 18.9 |
| Speed level 3 | 10.4 | 21.9 |
| Speed Turbo | 50.6 | 76.8 |

a Manufacturer provided information.

b Measured.

**Table A2. Calibration coefficients ^a^ of the optical particle counters (PurpleAir PA-II-SD).**

| **Monitor ID** | **Slope** | **R^2^** | **RMSE ^b^** |
| --- | --- | --- | --- |
| Monitor #1 | 0.43 | 0.99 | 272.61 |
| Monitor #2 | 1.11 | 0.99 | 461.40 |
| Monitor #3 | 1.24 | 0.99 | 248.83 |
| Monitor #4 | 1.15 | 0.99 | 197.85 |
| Monitor #5 | 1.58 | 0.97 | 896.25 |
| Monitor #6 | 1.27 | 0.99 | 486.15 |
| Monitor #7 | 1.32 | 0.99 | 192.03 |
| Monitor #8 | 1.21 | 0.99 | 486.16 |
| Monitor #9 | 1.32 | 0.99 | 456.67 |
| Monitor #10 | 0.45 | 0.99 | 450.60 |
| Monitor #11 | 1.25 | 0.99 | 607.67 |

^a^ The calibration model for each monitor was fitted in the form of $C_{TSI 3330}= {\beta_{0}+\beta}_{1}{\cdot C}_{PA}+\varepsilon$, where $C_{TSI 3330}$ is the particle count with the diameter ranging from 0.3 – 10 μm (#/cm^3^) measured by the reference instrument (TSI optical particle sizer model 3330, TSI Inc.); $C_{PA}$ is the raw particle count measured by the continuous optical particle counters (PurpleAir PA-II-SD; PurpleAir) in size bin >0.3 μm (#/cm^3^); $\beta_{0}$ is the intercept; $\beta_{1}$ is the slope; $\varepsilon$ is the residual. $\beta_{0}$ was set to zero for fitting.

^b^ RMSE: root mean square error. The RMSE of the post-calibrated optical particle counters were calculated using the equation $RMSE= \sqrt{\frac{\sum_{i=1}^{N} {({C_{TSI 3330} - C}_{PA_{cal}})}^{2}}{N}}$ where $N$ is the number of observations; $C_{TSI 3330}$ is the particle count with the diameter ranging from 0.3 – 10 μm (#/cm^3^) measured by the reference instrument (TSI optical particle sizer model 3330, TSI Inc.); ${C_{PA}}_{cal}$ is the post-calibrated particle count measured by the continuous optical particle counters (PurpleAir PA-II-SD; PurpleAir) in size bin >0.3 μm (#/cm^3^).

**Table A3. Summary of the total PAC working minutes monitored at each site.**

| **Site** | **Round** | **Number of PAC deployed** | **Total PAC working minutes monitored** | **Missing (%)** |
| --- | --- | --- | --- | --- |
| Site 1 | R1 | 13 | 256613 | 0.003 |
| Site 1 | R2 | 13 | 255824 | 0.31 |
| Site 1 | R3 | 13 | 256619 | 0.0004 |
| Site 2a | R1 | 10 | 197400 | 0 |
| Site 2a | R2 | 10 | 197400 | 0 |
| Site 2a | R3 | 10 | 197400 | 0 |
| Site 2b | R1 | 11 | 217140 | 0 |
| Site 2b | R2 | 11 | 217140 | 0 |
| Site 2b | R3 | 11 | 217140 | 0 |
| Site 3 | R1 | 3 | 59220 | 0 |
| Site 3 | R2 | 3 | 47979 | 18.9 |
| Site 3 | R3 | 3 | 59220 | 0 |
